# Decoding Neural Activity in Sulcal and White Matter Areas of the Brain to Accurately Predict Individual Finger Movement and Tactile Stimuli of the Human Hand

**DOI:** 10.1101/2021.04.06.21255006

**Authors:** Chad Bouton, Nikunj Bhagat, Santosh Chandrasekaran, Jose Herrero, Noah Markowitz, Elizabeth Espinal, Joo-won Kim, Richard Ramdeo, Junqian Xu, Matthew F. Glasser, Stephan Bickel, Ashesh Mehta

**Author notes:** Shared senior authorship (Chad Bouton, Ashesh Mehta).

## Abstract

Millions of people worldwide suffer motor or sensory impairment due to stroke, spinal cord injury, multiple sclerosis, traumatic brain injury, diabetes, and motor neuron diseases such as ALS (amyotrophic lateral sclerosis). A brain-computer interface (BCI), which links the brain directly to a computer, offers a new way to study the brain and potentially restore these losses in patients living with debilitating conditions. One of the challenges currently facing BCI technology, however, is how to minimize surgical risk. Minimally invasive techniques, such as stereoelectroencephalography (SEEG) have become more widely used in clinical applications since they can lead to fewer complications. SEEG electrodes also give access to sulcal and white matter areas of the brain but have not been widely studied in brain-computer interfaces. We therefore investigated the viability of using SEEG electrodes in a BCI for recording and decoding neural signals related to movement and the sense of touch and compared its performance to electrocorticography electrodes (ECoG) placed on gyri. Initial poor decoding performance and the observation that most neural modulation patterns were highly variable trial-to-trial and transient (significantly shorter than the sustained finger movements studied), led to the development of a feature selection method based on a repeatability metric using temporal correlation. An algorithm based on temporal correlation was developed to isolate features that consistently repeated (required for accurate decoding) and possessed information content related to movement or touch-related stimuli. We subsequently used these features, along with deep learning methods, to automatically classify various motor and sensory events for individual fingers with high accuracy. Repeating features were found in sulcal, gyral, and white matter areas and were predominantly phasic or phasic-tonic across a wide frequency range for both HD (high density) ECoG and SEEG recordings. These findings motivated the use of long short-term memory (LSTM) recurrent neural networks (RNNs) which are well-suited to handling both transient and sustained input features. Combining temporal correlation-based feature selection with LSTM yielded decoding accuracies of up to 92.04 +/-1.51% for hand movements, up to 91.69 +/-0.49% for individual finger movements, and up to 80.64 +/-1.64% for focal tactile stimuli to the finger pads and palm while using a relatively small number of SEEG electrodes. These findings may lead to a new class of minimally invasive brain-computer interface systems in the future, increasing its applicability to a wider variety of conditions.

## Introduction

There are currently 5 million people living with paralysis in the United States (Armour et al., 2016). The leading causes include stroke, spinal cord injury, multiple sclerosis, and amyotrophic lateral sclerosis (ALS). Brain-computer interface (BCI) technology was successfully demonstrated to form a neural bypass and restore volitional movement in paralysis during a first-in-human study that involved decoding movement-related neural signals recorded via microelectrodes implanted in motor cortex (Bouton et al., 2016). Furthermore, restoration of the sense of touch, important to regaining dexterous hand movement, has been demonstrated with BCI technology as well (Flesher et al., 2016). Microelectrodes and electrocorticography (ECoG) electrodes have been used for BCI applications, however, require a lengthy craniotomy to implant, thereby adding risk to the procedure. More recently, stereoelectroencephalographic (SEEG) electrodes have been used for mapping seizure origination in epileptic patients and is now widely accepted, offering a minimally invasive method for these procedures. Furthermore, adverse events associated with SEEG procedures occur at a significantly lower rate than with electrocorticography (ECoG) electrodes (Cardinale et al., 2013; Stricsek, Lang, & Wu, 2018). SEEG electrodes therefore may be a good alternative to ECoG or microelectrode arrays, which require a craniotomy to place, in brain computer interface (BCI) systems.

Decoding performance in BCI systems is an important consideration and can be directly impacted by the type of electrode used. ECoG and microelectrode arrays have been demonstrated in primates and humans for a variety of decoding applications including thought-controlled cursor movement and robotic arm control (Carmena et al., 2003; Hochberg et al., 2006). Decoding of individual finger movement has also been demonstrated in ECoG recordings (Kubánek et al., 2009). Very little work has been conducted to date, however, in assessing decoding performance when using SEEG electrodes for BCI applications. Basic two-dimensional cursor control has been demonstrated via SEEG electrodes (Vadera et al., 2013), in which the user wiggled their contralateral hand, or foot, to control the horizontal and vertical motion of a computer cursor respectively. Also, a BCI P300 Speller (single degree-of-freedom) was controlled through ECoG and SEEG electrodes implanted in and near the hippocampus (Shih & Krusienski, 2012)(Krusienski & Shih, 2011). Finally, grasp force related events were recorded and classified using SEEG electrodes recording from sulcal areas in motor cortex and from sensory cortex (Murphy et al., 2016).

One compelling application of SEEG electrodes is in a so-called bidirectional neural bypass for restoration of movement and the sense of touch. A minimally invasive approach is of particular interest since the bi-directional neural bypass application would require placement of electrodes in both motor and sensory areas of the brain. In fact, precise stimulation of sulcal and white matter areas for evoking focal percepts in the fingertips has also been demonstrated in humans recently using SEEG electrodes (Chandrasekaran et al., 2020). A bidirectional neural bypass needs to be effective in restoring a wide variety of movements including sustained movements to be useful for disabled users in activities of daily living. Previous work has shown that predominantly phasic (transient) neural modulation patterns were obtained during movements, but these studies were mostly centered on short or pulsed, but not sustained, movements (Chen et al., 2014; Flint et al., 2017; Shin et al., 2012). Also, recent work has further confirmed that predominantly phasic (transient) neural patterns with SEEG recordings during sustained grasping movements (Jiang et al., 2020). This suggests that using this type of activity as input features to a decoder for restoring sustained movement in an electronic neural bypass system may pose significant challenges. Another challenge stems from noise and drift that occurs in neuronal activity over time which can degrade decoding performance (Rule et al., 2020; Zhang et al., 2018). This emphasizes the importance of using effective signal processing and feature selection methods that extract reproducible features for decoding.

A number of strategies and different machine learning algorithms including linear classifiers, regression machines, support vector machines (SVMs), and deep neural networks have been used to decode neural signals recorded in the brain. In the handful of studies that have attempted to decode SEEG signals specifically, the results have been mixed. In one of these studies, three different hand gestures were decoded using SEEG signals with an accuracy of 78.70±4.01% (G. Li et al., 2017). In another study, SEEG electrodes placed in middle temporal regions led to fast typing of up to 14 characters/minute (D. Li et al., 2017). Lastly, another group decoded SEEG recordings from the auditory cortex and produced intelligible waveforms with 45-75% accuracy levels depending on the algorithm used (Akbari et al., 2019).

Here we investigated the temporal characteristics of neural signals for both motor and sensory events when recorded using SEEG electrodes and decoding methodologies that incorporated temporal correlation-based feature selection and deep learning methods. Specifically, we were interested in decoding sustained finger movements and evaluated long short-term memory (LSTM) based recurrent neural networks (RNNs), hypothesizing their memory cells and suitability to transient events may improve decoding accuracies (Hosman et al., 2019). Furthermore, with a long-term goal of restoring dexterous hand movement, we investigated decoding performance for individual finger movement and touch-related events at the fingertips. Our findings show that neural patterns recorded with SEEG electrodes are indeed mostly phasic in nature, but that LSTM-based deep learning networks combined with repeatability-based feature selection can produce high decoding accuracies.

## Methods and Materials

### Participants

Three patients voluntarily took part in this study that were undergoing pre-operative seizure monitoring for surgical treatment of intractable epilepsy. Functional magnetic resonance imaging (fMRI) was performed in participants 1 and 3 (P1 and P3) and implantation of either HD-ECoG grids and/or SEEG electrode leads was performed in each participant, and signals were recorded during various motor and sensory tasks (see Figure 1). The decisions on whether to implant, the electrode targets selected, and the duration for implantation were based entirely on clinical grounds without reference to this investigation. Some patients required re-implantation which is indicated by an underscore after the participant code (e.g. _02, _03). Patients were informed that participation in this study would not alter their clinical treatment, and that they could withdraw from the study at any time without jeopardizing their clinical care. All procedures and experiments were approved by the Northwell Institutional Review Board and participants provided informed consent prior to enrollment into the study.

**Figure 1.**
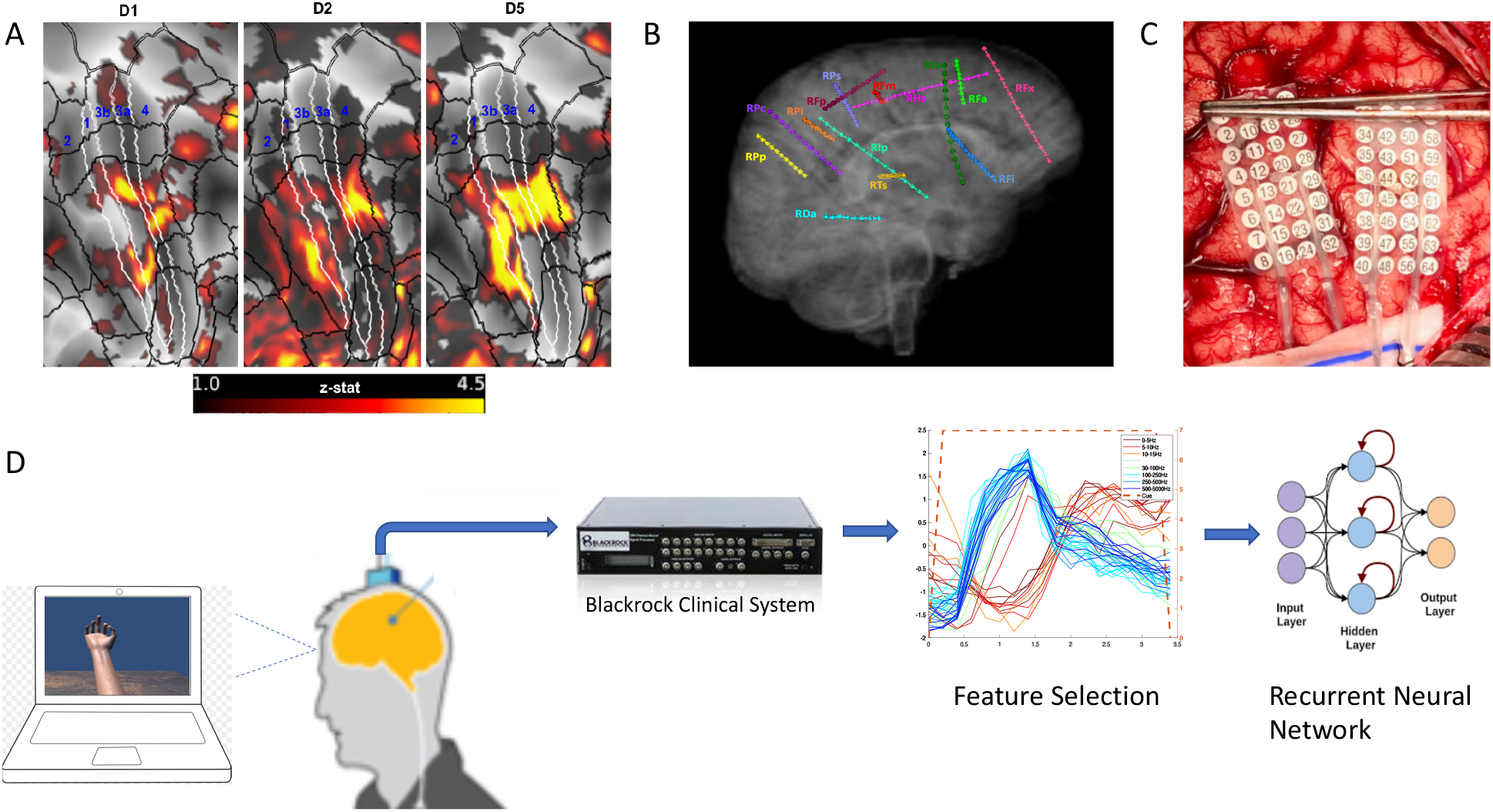
Functional magnetic resonance imaging and electrode placement. (A) Pre-surgical fMRI obtained while participant P1 pressed different buttons on a handheld device while watching videos showing desired movements. (B) Exemplary placement of SEEG electrodes (participant 3). (C) Photograph of HD ECoG electrode placement for recordings in participant P1_03. (D) Experimental setup where participants received visual cues of hand movements on a laptop computer with cues lasting 3 or 4s followed by a 3 or 4s period of rest; the clinical recording system (Natus Medical, Inc.) is not shown and was always connected for continuous data acquisition.

### Imaging

Participants were scanned on a 3T MRI scanner (Skyra, Siemens, Germany) with a 32-channel head coil. Human Connectome Project (HCP)-like structural and functional MRI were acquired: T1-weighted (T1w) 3D MPRAGE sequence, 0.8 mm isotropic resolution, TR/TE/TI = 2400/2.07/1000 ms, flip angle = 8 degree, in-plane under-sampling (GRAPPA) = 2, acquisition time 7 min; T2-weighted (T2w) 3D turbo spin echo (SPACE) sequence, 0.8 mm isotropic resolution, in-plane under-sampling (GRAPPA) = 2, TR/TE = 3200/564 ms, acquisition time 6.75 min; task fMRI using the CMRR implementation of multiband gradient echo echo-planar imaging (EPI) sequence (Setsompop et al., 2012), 2.1 mm isotropic resolution, 70 slices with a multiband factor of 7 (Xu et al., 2013), FOV 228 mm × 228 mm, matrix size 108 × 108, phase partial Fourier 7/8, TR/TE = 1000/35 ms, flip angle = 60 degree, phase encoding direction = anterior-posterior (A-P), echo spacing = 0.68 ms, 240 volumes in 4 min; and a pair of reversed polarity (A-P / P-A) spin echo EPI field mapping acquisitions with matched echo train length and echo spacing to the fMRI acquisition. The task was button-pressing on the PST button response unit (Psychology Software Tools, Sharpsburg, PA, USA) using a single finger (wrist restrained with strap on the button response unit and neighboring fingers taped down with medical tape), repeating 6 times of 20-second off (resting with cue of a blank dark screen) and 20-second on (tapping with continuous video cue of the same finger motion presented from a projector screen). Participant P1 performed three repetitions of the task for each of thumb, index, and little fingers (phase-encoding direction A->P) while participant P3 performed two repetitions for each of thumb, index, and middle fingers (phase-encoding directions A->P and P->A). The MRI preprocessing began with the HCP minimal preprocessing pipelines version 3.27 (Glasser et al., 2013) including, motion correction, distortion correction, cortical surface reconstruction and subcortical segmentation, generation of T1w/T2w-based myelin content and cortical thickness maps, transformation of the fMRI data to MNI and CIFTI grayordinate standard spaces using folding-based registration with MSMSulc, and 2 mm full-width half maximum (FWHM) surface and subcortical parcel constrained smoothing for regularization. The fMRI data were cleaned of spatially specific structured noise using the HCP’s multi-run (version 4.0) ICA-FIX for multi fMRI (multiple finger tasks) and linear trends without regressing out motion parameters. Somatotopic functional responses were estimated (first-level for participant P1 and second-level fixed-effect averaging of the two phase-encoding directions for participant P3) using a generalized linear model (GLM)-based fMRI analysis (Woolrich et al., 2001) on the grayordinate data space for each finger.

### Electrode localization

The SEEG electrodes (PMT Corporation, Chanhassen, MN, USA) consisted of 16 contacts, cylinders with 2 mm length, 0.8 mm diameter, and 4.43 mm spacing (center to center). The HD-ECoG grids (PMT Corporation) consisted of 2 mm diameter flat contacts, in participant P1 with an 8×8 arrangement with 5 mm spacing (center to center) and in participant 2 with 16×16 contact arrangement with 4 mm spacing. Since both patients had clinical indications that required mapping of the sensorimotor cortex, task-based fMRI activation maps were used to guide electrode placement. For digital localization of the electrodes, we used the freely available iElvis toolbox, available at https://github.com/iELVis/ (Groppe et al., 2017). Briefly, the electrodes were manually localized using the software BioImage Suite (http://www.bioimagesuite.org) on a postimplant CT which was co-registered using an affine transformation (6 degrees-of-freedom FLIRT; www.fmrib.ox.ac.uk/fsl) to the pre-implantation 3T high-resolution T1w MRI. We used the FreeSurfer output from the HCP minimal processing pipeline (Glasser et al., 2013) to obtain the pial surface. The subdural HD-ECoG electrodes were projected to the smoothed pial surface. The smoothed pial surface, also called the outer smoothed surface, is generated by Freesurfer and wraps tightly around the gyral surfaces of the pial layer while bridging over the sulci. No correction was applied to SEEG electrode coordinates. To visualize the fMRI activation maps and the electrodes simultaneously, we used HCP Connectome Workbench (http://www.humanconnectome.org/software/connectome-workbench). Before importing the electrode coordinates into Connectome Workbench, we applied a RAS coordinate offset as follows – *transformed_RAS_coordinates = Norig*inv(Torig)*RAS_coordinates* where the transformation matrices *Norig* is obtained by *mri_info --vox2ras [subject]/mri/orig*.*mgz* and *Torig* is obtained by *mri_info --vox2ras-tkr [subject]/mri/orig*.*mgz* The transformed coordinates were then imported as foci using the cortical surfaces into Connectome Workbench.

### Recording of neural activity

In addition to the clinical recording system, neural activity was recorded using a Neuroport System (Blackrock Microsystems, Salt Lake, Utah) with a sampling rate of 10 kHz while participants performed the various tasks. These tasks included hand/finger movements and mechanical stimulation of the fingertips of their hand using a von Frey filament (TouchTest Sensory Probes) of evaluator size, 3.61 (0.4 g). An electrode located in soft tissue lacking neural activity was used as the system ground. Subsequent analysis involved multiple steps to extract information regarding power modulation in different frequency bands. Signals from neighboring electrodes were subtracted in software to provide bipolar data with reduced noise. Non-overlapping Blackman windows of 200 ms in length were applied to the data, followed by a short Fast Fourier Transform (sFFT) for each window (with a resulting frequency resolution of 5 Hz). The amplitude information at each frequency was then integrated across pre-selected frequency bands as follows: 0–10, 10–15, 15–30, 30–100, 100–500, and 500–5000 Hz. These integrated amplitude features (IAFs) for all bipolar recordings were standardized by subtracting their mean and dividing by their standard deviation across the entire task.

### Decoding and feature selection methods

Epochs aligned with each visual cue (animated hand) presented to the participant during each finger movement were created, starting at the cue onset, and extending to 400 ms after the cue offset. All cue-aligned trials for each IAF were averaged to form a composite temporal response. To quantify the repeatability of potential features, an algorithm based on temporal correlation was used to compute the mean correlation coefficient (MCC) by averaging the correlation coefficients obtained for temporal responses for each trial with respect to the composite temporal response. Features were then selected based on their MCC value. The range of the MCC values chosen were 0.4 to 0.6, known as moderate correlation levels, as they led to improved decoding performance. MCC values outside this range tended to worsen the decoding performance. Selected features were then used to train an LSTM type RNN using Matlab and the Deep Learning Toolbox (R2019b, Update 4). Training parameters are as noted in Results section.

## Results

Functional MRI images were obtained in participants P1 and P3 and post-surgical CT images in all participants. The fMRI images for the button press paradigm are shown in Figure 2, along with post-surgical CT images highlighting electrodes having MCC values greater than 0.6 (moderate correleation) during finger flexion tasks. Furthermore, during the fMRI the peak activity was primarily located in the central sulcus for all three digits during the task.

**Figure 2.**
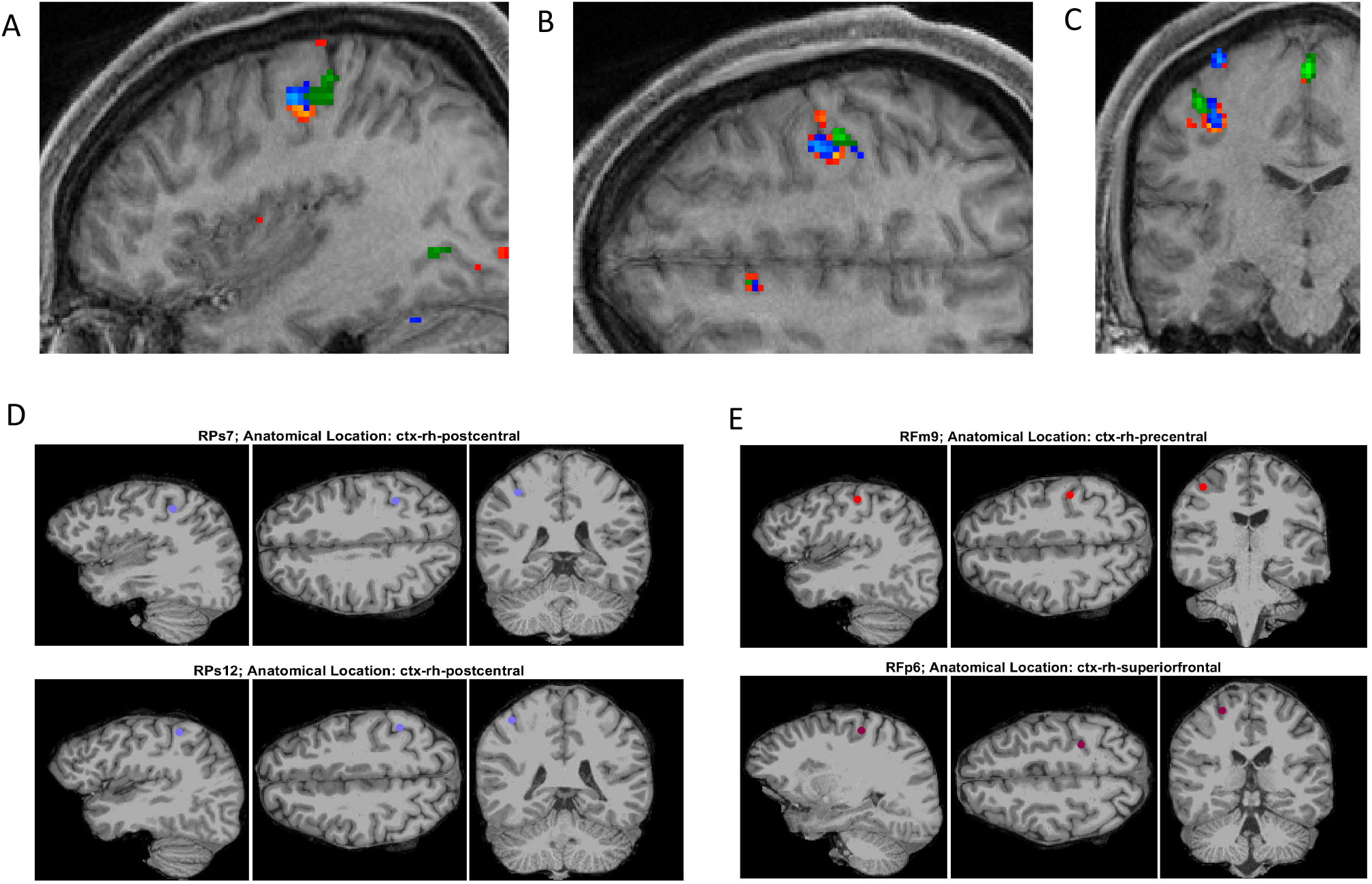
Functional MRI and post-surgery CT images for participant P3 related to finger tasks.(A-C) Functional MRI images in orthogonal planes (sagittal, transverse, and coronal) obtained during button press task for digits 1 (orange-red), 2 (blue), and 3 (green). (D and E) Post-surgical CT images of SEEG electrode sites found to have a repeatability metric (MCC) value greater than 0.6 (for time range of 0-4.4s with respect to cue onset) for flexion of digits 1 and 3.

During the hand and finger movement tasks, phasic (transient) and phasic-tonic (transient-sustained) evoked responses were identified, using temporal correlation analysis, in all frequency bands analyzed across the 0 to 5000Hz range (as shown in Figure 3). As expected, most evoked responses in the delta, theta, alpha/mu, and beta bands exhibited a decrease in amplitude after visual cue presentation to the participant, whereas an increase in power was observed in the gamma bands. An exception was observed in one participant (P3) where both increases and decreases were observed in the lower frequency bands.

**Figure 3.**
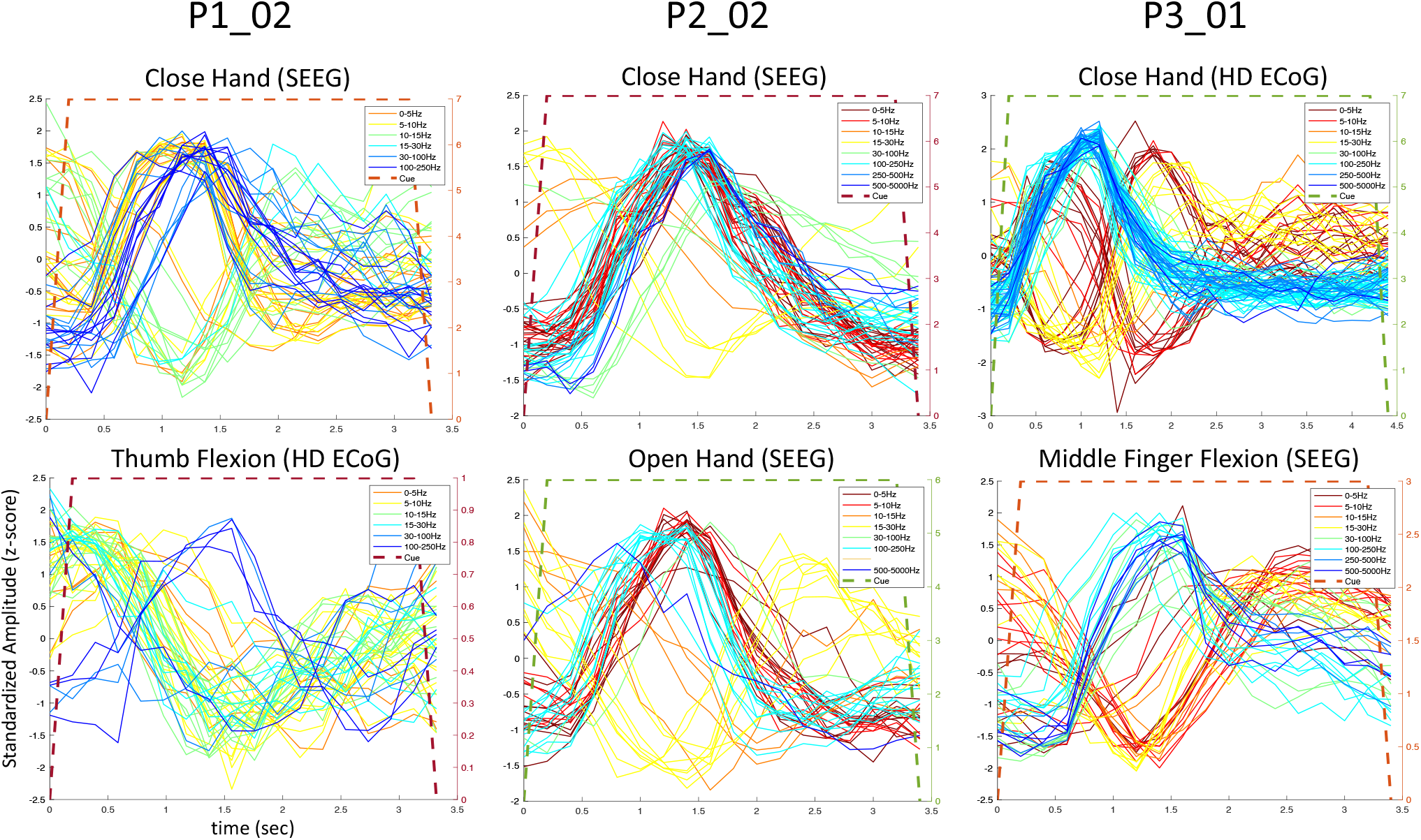
Predominantly phasic neural activity for sustained hand and finger movements in SEEG and ECoG recordings. Representative set of composite temporal responses across three study participants and different hand and finger movements are shown. In both the SEEG (depth) and HD ECoG (surface) recordings, the responses across a wide range of frequency bands are predominantly phasic in nature. The MCC (repeatability metric) was greater than 0.6 for all plots in this figure except in P1_02 where it was 0.5 for ‘Close hand (SEEG)’ and 0.4 for ‘Thumb Flexion (HD ECoG).’

To further examine the transient nature of the evoked responses, an analysis extended to time periods after both the visual cue onset and offset was performed. As shown in Figure 4, transient responses in SEEG electrodes were observed after the cue offset as well, and in both motor and sensory tasks in participant P3. In the sensory tasks, mechanical tactile stimuli (rapid tapping) were presented throughout the entire cue period at the fingertips (center of pads) and palm areas.

**Figure 4.**
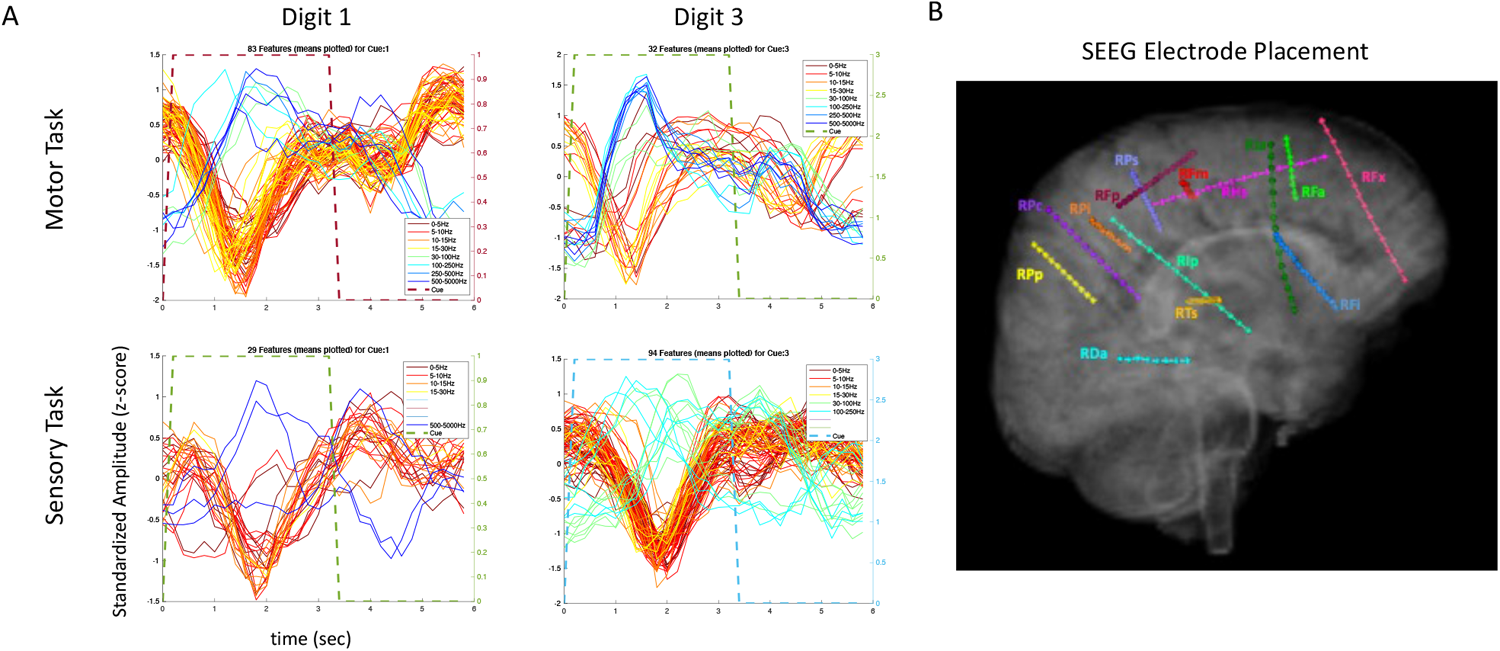
Amplitude modulation and electrode placement in P3_01 (SEEG electrodes). (A) Composite temporal patterns of amplitude modulation in various frequency bands for motor and sensory tasks. (B) Electrode placement in various regions of the brain.

**Figure 5.**
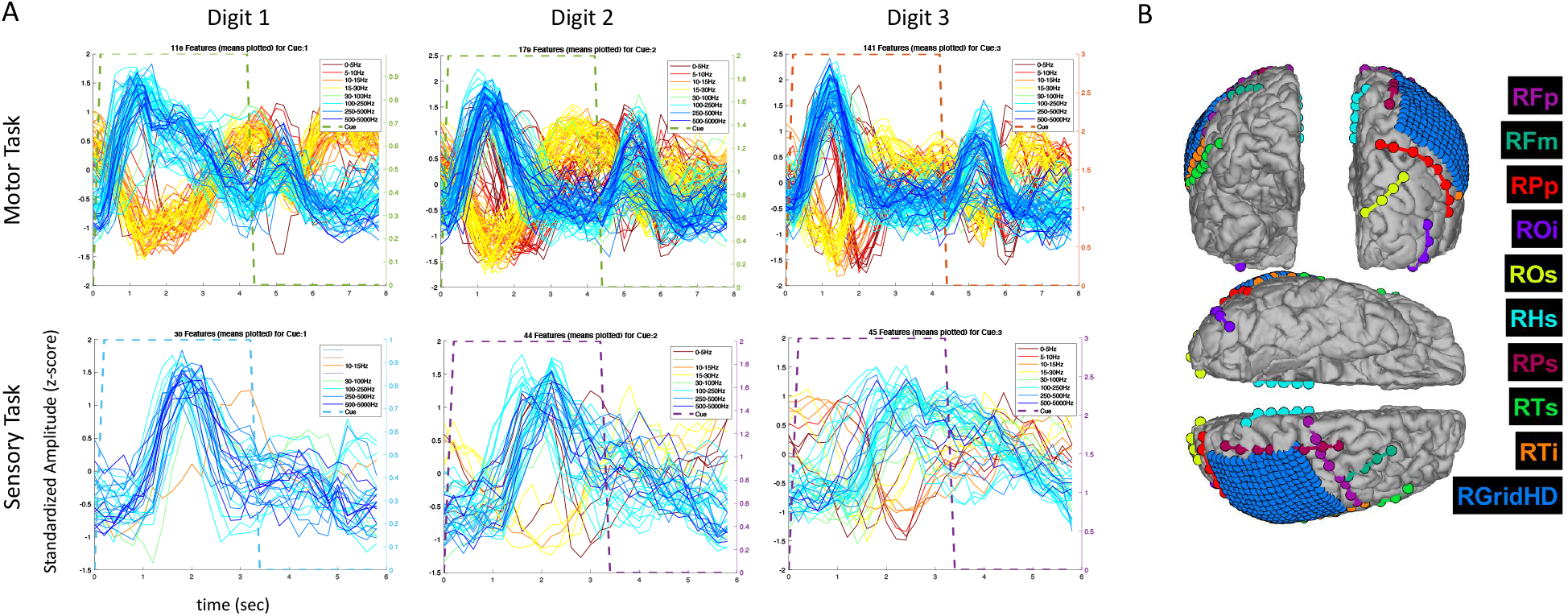
Amplitude modulation and electrode placement in P3 (ECoG and strip electrodes). (A) Averaged (across trials) temporal patterns of amplitude modulation in various frequency bands for motor and sensory tasks. (B) Electrode placement in various surface regions of the brain.

The extended time analysis was performed in the same participant (P3) for HD ECoG recordings during both motor and sensory tasks. Similar phasic and phasic-tonic responses were observed but more features with high temporal correlation were identified (as the HD ECoG array was placed directly over the sensorimotor cortex). Interestingly, responses with more tonicity were observed on some digits, but which digit differed depending on whether a motor or sensory task was being performed.

Given that the evoked responses observed were primarily phasic in nature and varied in their temporal signatures and stable features (features with high repeatability) are desirable for accurate decoding, temporal correlation analysis was introduced to quantify repeatability of potential features. It was hypothesized using temporal correlation-based feature selection would improve decoding performance, particularly since many of the SEEG electrodes and some of the HD ECoG electrodes were not located in areas believed to be related to movement or sensation.

Several motor tasks were first implemented where the participant performed sustained hand and finger movements including ‘open hand’ (wide opening of hand with splayed fingers), ‘close hand ‘(make a fist), and finger flexion (sustained flexion of individual fingers). As shown in Figure 6, using temporal correlation-based feature selection significantly improves decoding accuracy for both SVM and LSTM type algorithms when using SEEG or HD ECoG type electrode recordings in participant P3. This trend was also observed in the other participants. Note the task used for the HD ECoG data sets involves more finger movements (three instead of two), however the same improvement in accuracy is observed when temporal correlation-based feature selection is used. Ultimately, in both SEEG and HD ECoG type recordings, the highest accuracy is obtained using temporal correlation-based feature selection with a LSTM type algorithm.

**Figure 6.**
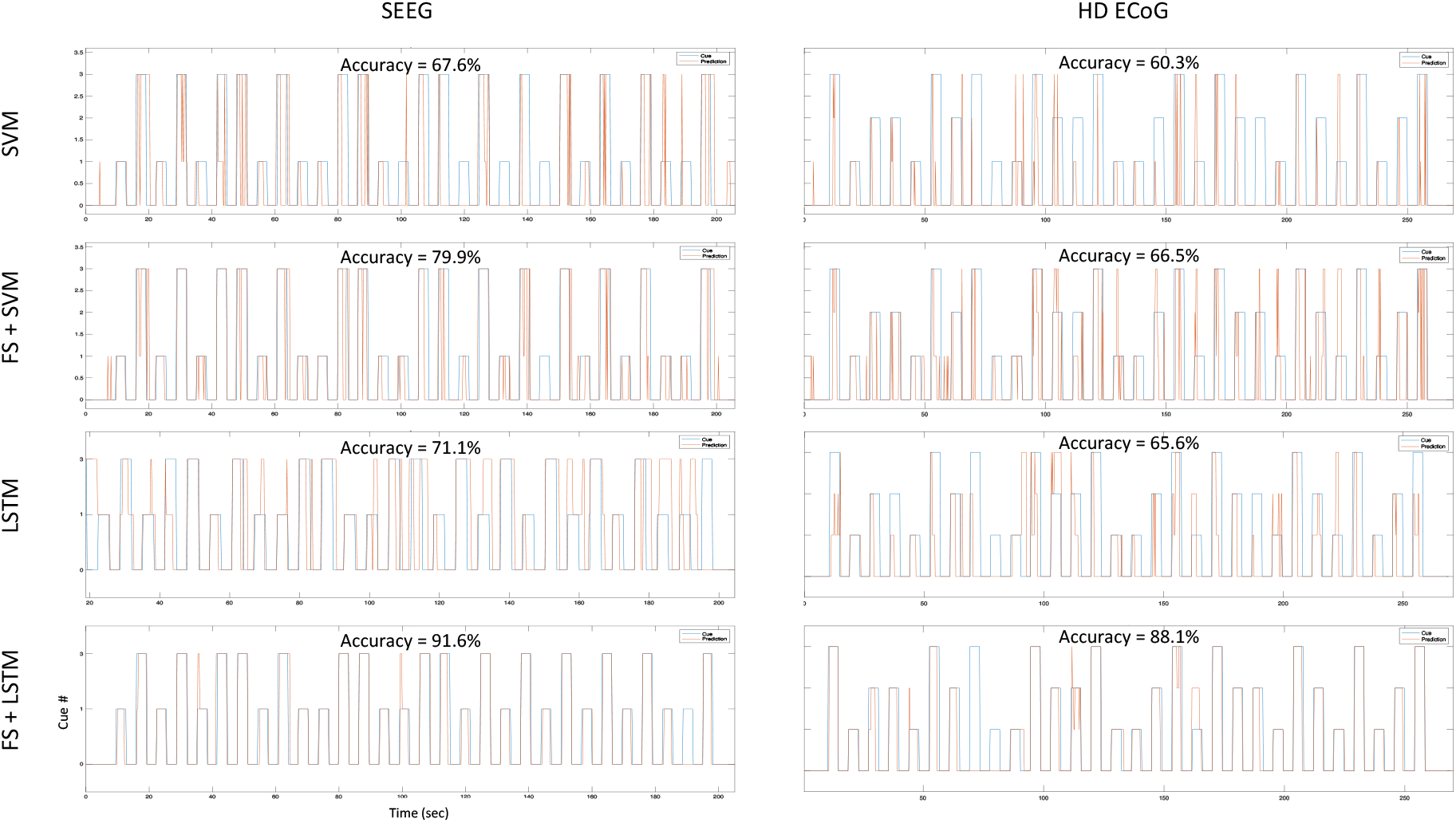
Decoding accuracies for different architectures during thumb, index (HD ECoG only), and middle finger flexion task. Independent test sets results are shown for four different decoding architectures tested offline with SEEG and ECoG recordings in participant P3: the first row results are for a multi-class nonlinear SVM using a gaussian kernel without feature selection, the second row are results for an architecture that performs repeatability-based feature selection (MCC > 0.6) and SVM, the third row shows results for a LSTM recurrent neural network without feature selection, and the last row shows results for an architecture using feature selection as described and a LSTM recurrent neural network (with 400 hidden units).

In Figure 7 the electrode locations for all three participants (red=P1, green=P2, and blue=P3), mapped to a standard glass brain based on the Yeo 7 atlas, are shown for the finger flexion task involving thumb flexion (+) and middle finger flexion (*) prompted by visual cues. Temporal correlation analysis was performed and electrodes that yielded a MCC value greater than 0.6 are shown using a colored symbol and a diamond shape is used to denote that the MCC value was greater than 0.6 for a given electrode during *both* the thumb and middle flexion cued movement periods. Overall, most of the electrodes with significant MCC values are located in the sensorimotor areas for all three participants.

**Figure 7.**
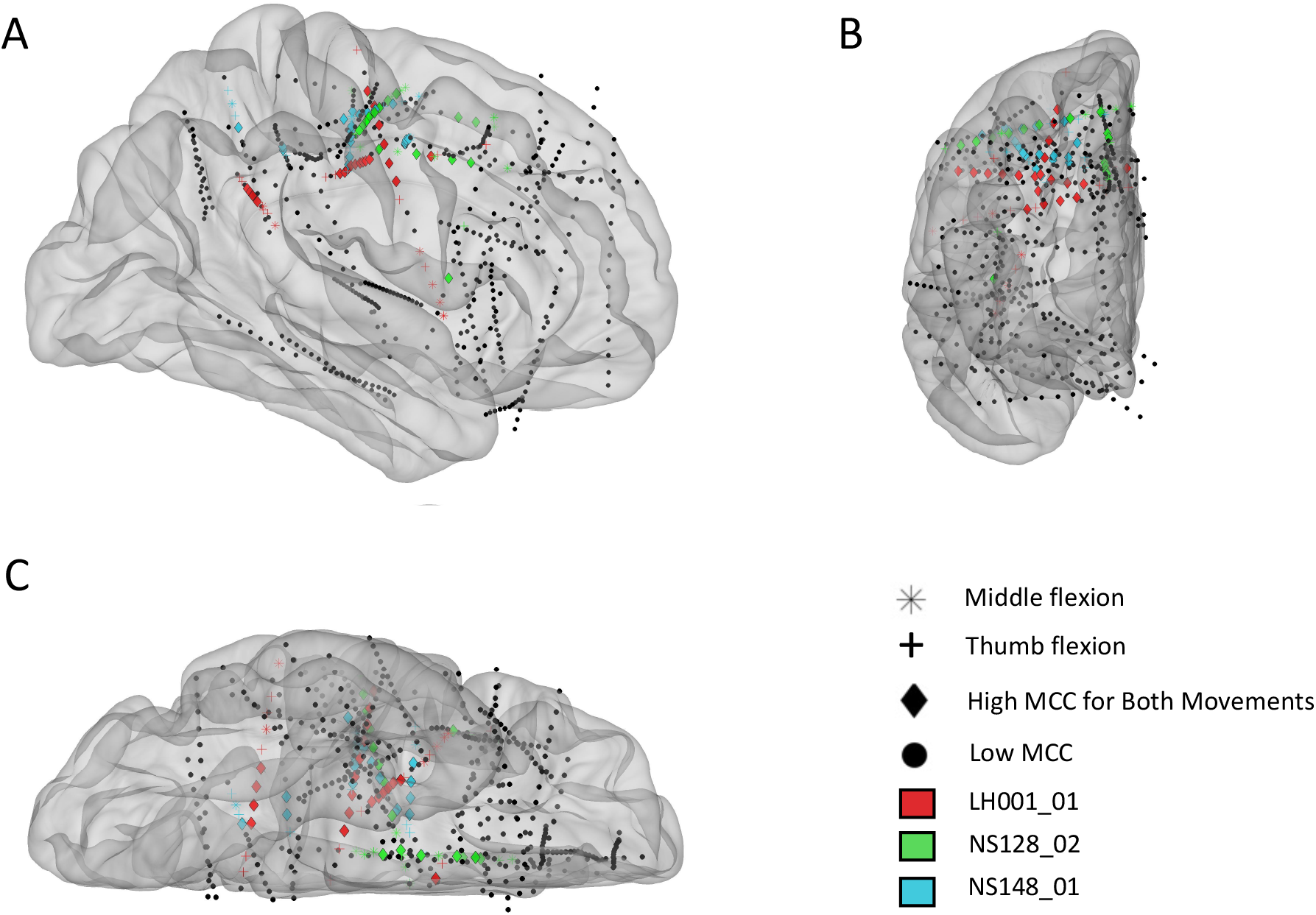
Electrode locations (right hemisphere) mapped to standard brain based on the Yeo 7 atlas for participants P1_01 (red), P2_02 (green), and P3_01 (blue). (A) Sagittal view from lateral perspective. (B) Coronal view from anterior perspective. (C) Transverse view from inferior perspective. The colored electrodes had high temporal correlation (MCC>0.6) during a cued finger task including sustained thumb and middle flexion movements, separately, and the symbols mark which movement(s) had high MCC values: thumb flexion (cross), middle finger flexion (asterisk), or both (diamond).

To further examine the location of electrodes with high temporal correlation, a functional network map was created. Electrodes within 3mm of the cortical surface were snapped to the nearest point on the surface of the brain and shown on an inflated standard brain based on the Yeo 7 network atlas. As shown in Figure 8, most of the electrodes with high temporal correlation (MCC>0.6) are located in the somatomotor, dorsal/ventral attention, and frontoparietal regions. Only two electrodes in the second subject (P2/green) have a high MCC value, but these are the only two electrodes implanted in this region (as confirmed by the lack of spherical green symbols).

**Figure 8.**
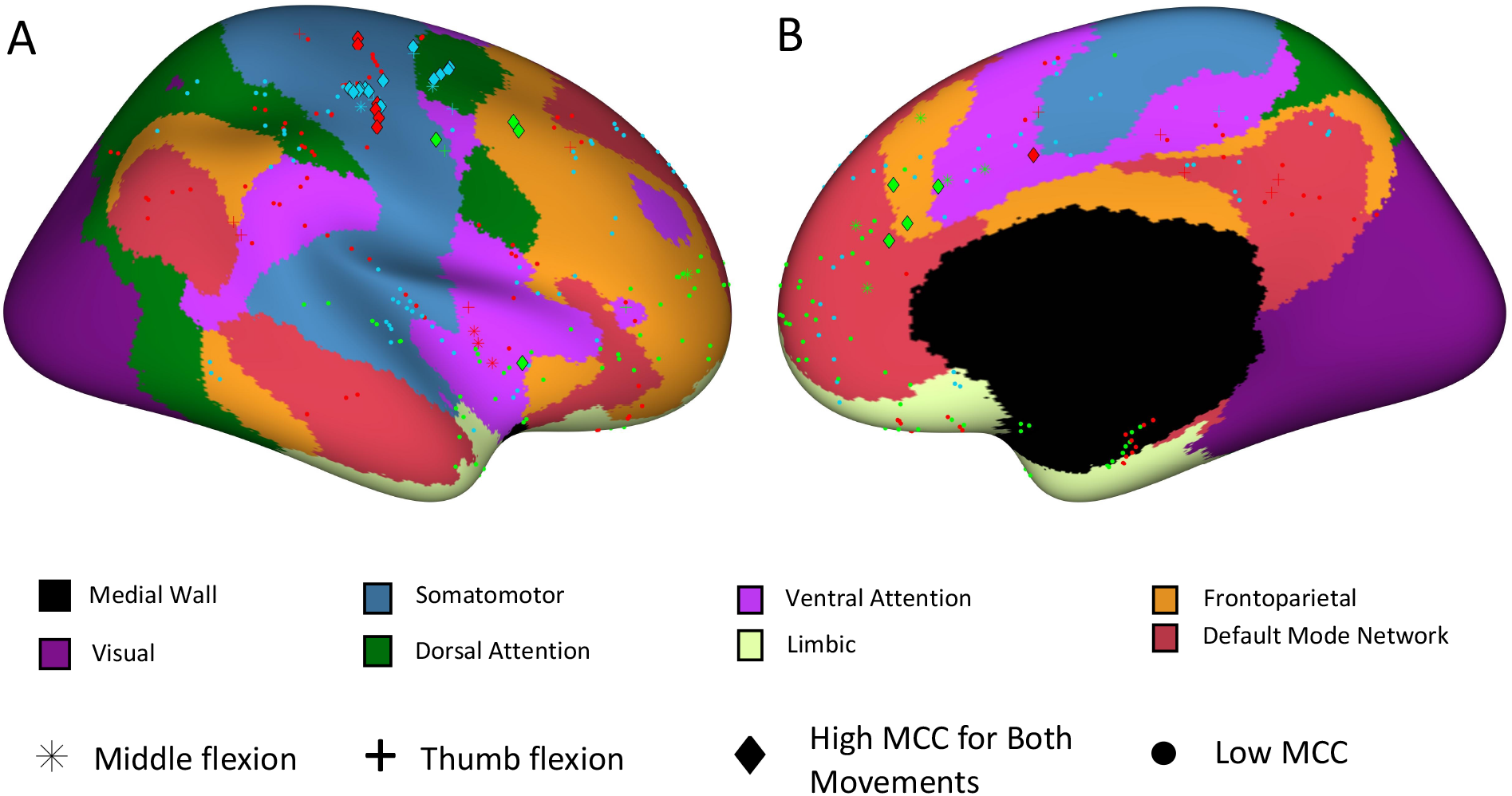
Electrode site locations with respect to functional networks for participants P1_01 (red), P2_02 (green), and P3_01 (blue). (A) Sagittal view from lateral perspective. (B) Sagittal view from medial perspective.

Decoding results are shown in Table 1 for motor tasks where the participant performed sustained hand and finger movements including open (wide open of hand with splayed fingers), close hand (make a fist), and finger flexion (sustained flexion of individual fingers). The highest decoding accuracy was achieved in SEEG recordings for open-close hand and thumb-middle finger flexion tasks, however, the HD-ECoG recording for the finger flexion task involved more classified states. Another interesting finding was that many SEEG electrode sites with high temporal correlation (MCC>0.6), and therefore selected for decoding, were found to be in white matter. In fact, for the open-close hand task, over fifty percent of these sites were in white matter for two of the three participants (16 of 27 sites for P2_02, 9 of 16 sites for P3_01, and 3 of 19 sites for P1_03).

**Table 1.**
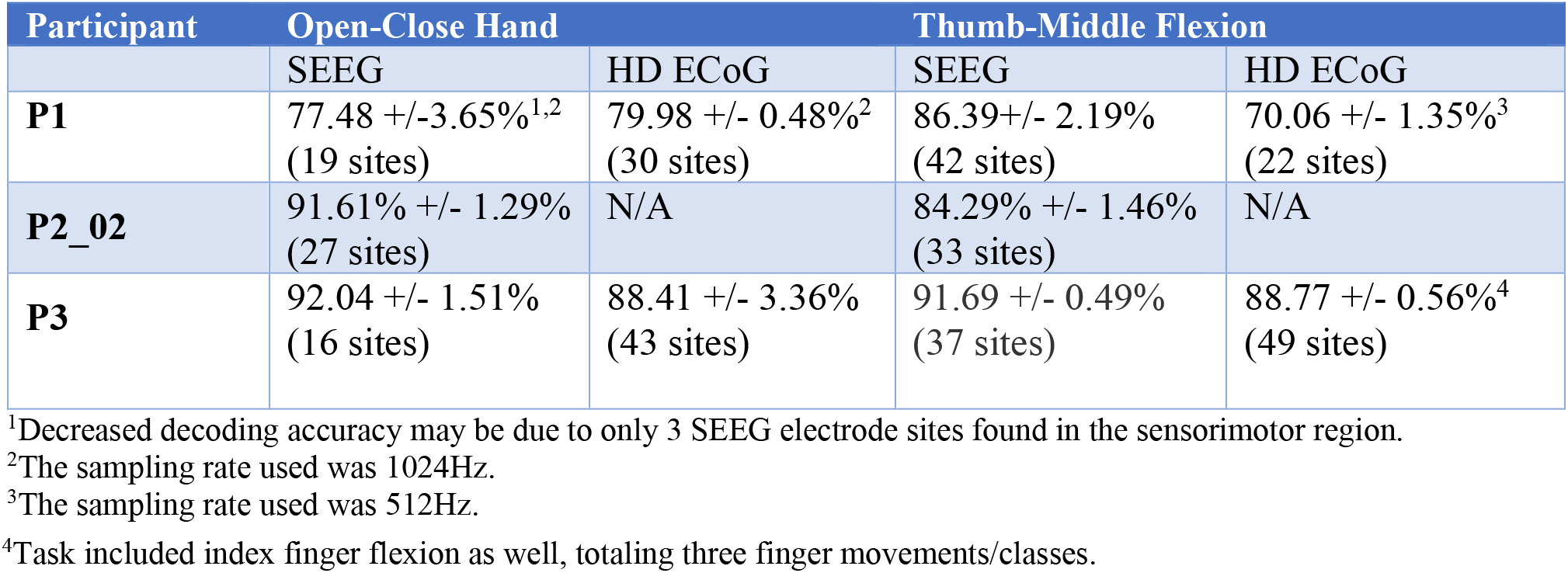
Decoding performance (mean +/-SD) for sustained hand movements when using a recurrent neural network (LSTM type) with repeatability-based feature selection. MCC threshold ranged from 0.55 to 0.6, the number of hidden units ranged from 400 to 600, and the patience ranged from 2 to 7.

**Table 2.**
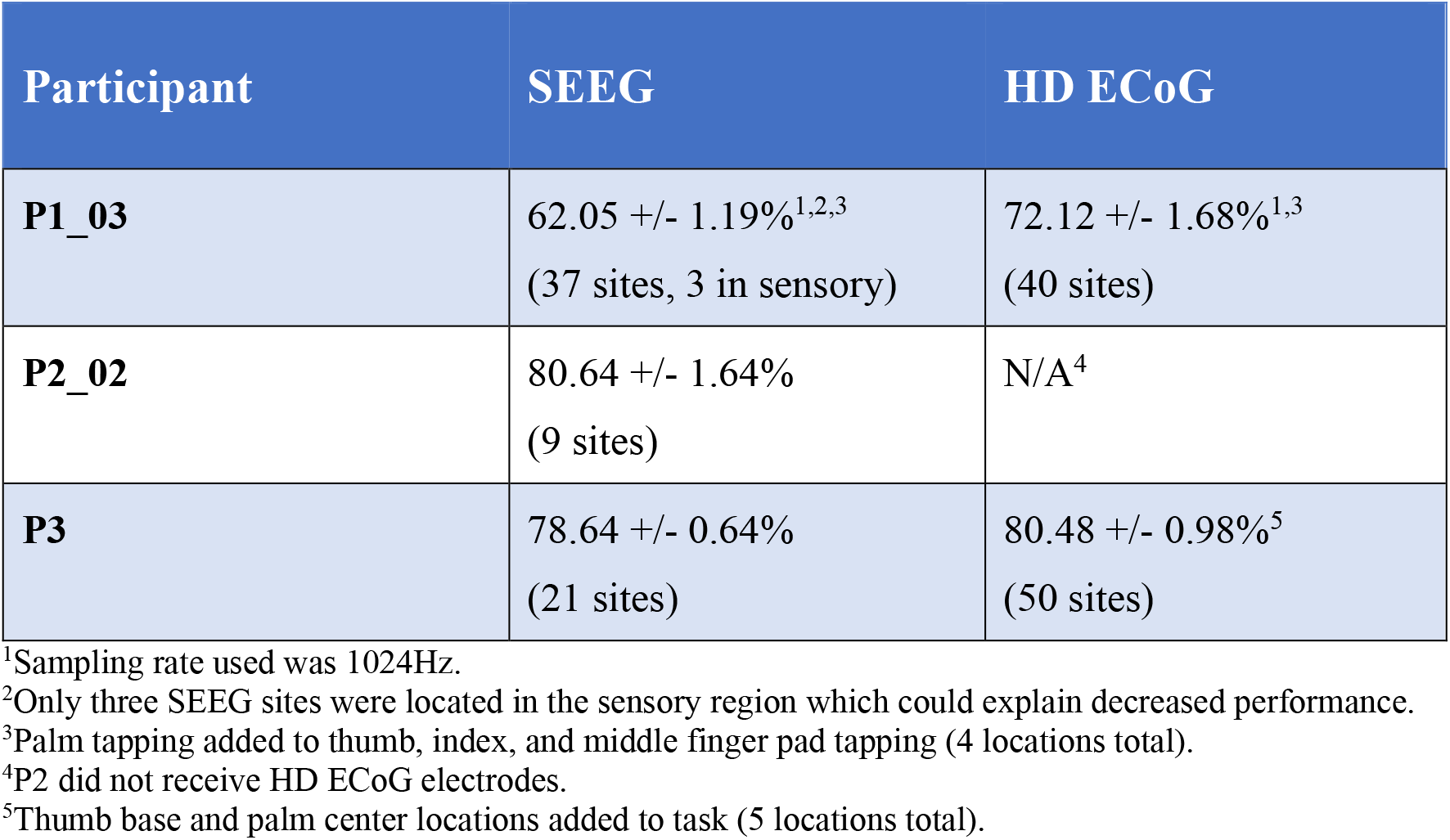
Decoding performance (mean +/-SD) for tactile stimuli (tapping) applied to thumb, index, and middle finger pads (and sometimes base of thumb or center of palm) when using the LSTM with temporal correlation-based feature selection algorithm.

During the tactile stimuli task where a mechanical filament was used to repeatedly tap the pads of various fingers, and sometimes palm areas, decoding results were obtained for SEEG and/or HD ECoG recordings. The decoding accuracies for these different recording modalities and the three different participants are shown in Table 1. The decoding accuracy results (when using an LSTM based algorithm with temporal correlation-based feature selection) for SEEG recordings were higher than those obtained with HD ECoG recordings. However, the HD ECoG tasks included more stimuli locations, therefore making it difficult to do a direct comparison.

## Discussion

In this study we showed minimally invasive SEEG electrodes can record stable and information-rich neural signals related to movement and tactile sensation. Using a temporal correlation-based feature selection method, we identified and extracted repeating neural patterns that consistently occurred during motor and sensory events. These features were used as inputs to a LSTM type recurrent neural network which was able to reliably and accurately predict finger movement and focal tactile stimuli presented at the pads of the fingers.

High decoding accuracy was demonstrated while using SEEG methods in multiple participants across multiple tasks and shown to be comparable to the decoding performances achieved with HD ECoG. This was unexpected given the relatively low spatial resolution of SEEG electrodes (4.43mm site spacing) and that there were fewer electrodes placed in the sensorimotor area as compared to the HD-ECoG grid electrodes. Furthermore, very few studies have demonstrated the application of SEEG in neural decoding and in the BCI field. In fact, the authors are not aware of any other previous study demonstrating the use of SEEG/depth electrodes for neural decoding of both motor and sensory stimuli events.

This study had limitations including the number of participants and task variation. In the future, expanding the study to include additional participants will allow further mapping of sulcal and white matter areas which are less charted than the gyri. In the current study reported here, not all tasks were completed for all participants due to session time constraints, therefore direct comparison of decoding performance for all tasks was not possible. Despite this, the data suggest SEEG recordings, combined with the methods presented, can achieve high decoding accuracies using relatively few electrode sites.

With the clear advantage of being minimally invasive, SEEG electrodes may be a viable option for use in brain-computer interface systems and specifically in a bidirectional neural bypass for paralysis applications. Although SEEG electrodes have been limited to acute use, similarly constructed depth electrodes, such as those used in deep brain stimulation for Parkinson’s disease, which have been implanted for multiple years, and some investigational devices have recording electrodes such as the Summit™ RC+S developed by Medtronic (Stanslaski et al., 2018).

The temporal correlation feature selection method was also found to identify locations within the brain involved in processing sensory stimuli that corresponded to the locations identified through cortical stimulation methods (Chandrasekaran et al., 2020). Passive methods such as this that do not involve electrical stimulation are attractive for mapping procedures in epileptic patients as they reduce the risk of inducing a seizure. The methods demonstrated in this study have many potential applications in mapping, movement and sensory restoration, and augmentative communication for patients living with paralysis, sensory loss, ALS, epilepsy, brain injury, and many other neurological conditions.

## Conclusion

Here we explored the viability of using minimally invasive SEEG methods and electrodes for decoding neural activity related to motor and sensory events. As observed in ECoG recordings, neural signals produced by SEEG electrodes are primarily phasic (transient) in nature, even during sustained motor tasks. When a feature selection method based on temporal correlation (a measure of feature repeatability) was implemented, decoding accuracy increased with both SVM and LSTM type recurrent neural network approaches. Furthermore, the overall decoding accuracy for SEEG recordings was comparable to the performance observed in ECoG recordings. Our findings support that SEEG can be an effective approach for neural decoding and for use in brain-computer interface systems. Finally, this minimally invasive approach reduces risk and may become the preferred approach for many BCI applications including restoration of movement and tactile sensation in impaired users.

## Data Availability

Data will be provided upon written request and approval.

## Acknowledgements

The authors would like to thank Todd Levy for his work and support on the graphical user interface and related software development. The authors would also like to thank Kevin J. Tracey for his insights, guidance, and for reviewing this manuscript.

## Author Contributions

SC, SB, CEB, and ADM designed the study. JWK and JX designed and performed the fMRI procedures and analyzed the data. ADM performed the SEEG leads and HD-ECoG grid implantations. NM and EE digitized and co-registered the electrode locations. SC, SB, JH, NAB, RR, and CEB performed all the experiments. CEB and SC analyzed data from these experiments. MFG and JX provided key insights into cortical anatomy, help using the workbench software and generating relevant figures for the manuscript. All authors contributed towards interpreting the results of the experiments. CEB wrote the initial draft of the paper and all authors provided critical review, edits and approval of the final manuscript.

